# The clinical course of COVID-19 in the outpatient setting: a prospective cohort study

**DOI:** 10.1101/2020.09.01.20184937

**Authors:** Paul W. Blair, Diane Brown, Minyoung Jang, Annukka A.R. Antar, Jeanne C. Keruly, Vismaya S. Bachu, Jennifer L. Townsend, Jeffrey Tornheim, Sara C. Keller, Lauren Sauer, David L. Thomas, Yukari C. Manabe, on behalf of the Ambulatory COVID Study Team

## Abstract

**Background:** Outpatient COVID-19 has been insufficiently characterized.

**Objective:** To determine the progression of disease and subsequent determinants of hospitalization.

**Design:** A prospective outpatient cohort.

**Setting:** Outpatients were recruited by phone between April 21 to June 23, 2020 after receiving outpatient or emergency department testing within a large health network in Maryland, USA.

**Participants:** Outpatient adults with positive RT-PCR results for SARS-CoV-2.

**Measurements:** Symptoms, portable pulse oximeter oxygen saturation (SaO_2_), heart rate, and temperature were collected by participants on days 0, 3, 7, 14, 21, and 28 after enrollment. Baseline demographics, comorbid conditions were evaluated for risk of subsequent hospitalization using negative binomial, logistic, and random effects logistic regression.

**Results:** Among 118 SARS-CoV-2 infected outpatients, the median age was 56.0 years (IQR, 50.0 to 63.0) and 50 (42.4%) were male. Among those reporting active symptoms, the most common symptoms during the first week since symptom onset included weakness/fatigue (67.3%), cough (58.0%), headache (43.8%), and sore throat (34.8%). Participants returned to their usual health a median of 20 days (IQR, 13 to 38) from the symptom onset, and only 65.5% of respondents were at their usual health during the fourth week of illness. Over 28 days, 10.9% presented to the emergency department and 7.6% required hospitalization. Individuals at the same duration of illness had a 6.1 times increased adjusted odds of subsequent hospitalization per every percent decrease in home SaO_2_ (95% confidence interval [CI]: 1.41 to 31.23, p=0.02).

**Limitations:** Severity and duration of illness may differ in a younger population.

**Conclusion:** Symptoms often persisted but uncommonly progressed to hospitalization. Home SaO_2_ might be an important adjunctive tool to identify progression of COVID-19.

**Registration:** Clinicaltrials.gov NCT number: NCT04496466

**Funding Source:** The Sherrilyn and Ken Fisher Center for Environmental Infectious Diseases Discovery Program and the Johns Hopkins University School of Medicine

## INTRODUCTION

SARS-CoV-2 (severe acute respiratory syndrome coronavirus 2) is the cause of the COVID-19 pandemic that has affected nearly every region of the world and by August 8^th^, 2020 is responsible for the deaths of more than 723,000 people(1). In persons who are hospitalized, the clinical features of COVID-19 and disease course are well described(2-4). However, most SARS-CoV-2 infected persons are not hospitalized, and relatively little is known about the progression of symptoms, clinical outcomes, and severity predictors among outpatients(5-7). The prevalence and time course of unique clinical features of COVID-19 such as the occurrence of low oxygen saturations with a delayed patient sense of dyspnea, or ‘silent hypoxemia,’ has yet to be fully characterized(8-10). Additionally, seroprevalence studies suggest that the number of outpatient cases are much greater than have been reported(11).

To investigate COVID-19 in the home setting, a prospective outpatient observational cohort was recruited and studied using structured measurements to characterize the course of disease. To better study risk factors for severe disease, the cohort recruitment efforts enriched for older individuals (12). Given the dominance of pulmonary syndromes in those hospitalized and to investigate the incidence of asymptomatic hypoxemia, we supplemented home monitoring with daily pulse oximetry(9).

## METHODS

### Study design

In an Institutional Review Board-approved study, persons ≥ age 18 who attended one of the Johns Hopkins Health System COVID-19 testing sites and tested positive for SARS-CoV-2 were offered enrollment in the study, excluding patients hospitalized at the time of screening. To maximize recruitment of older persons, we focused recruitment to individuals who were midlife adults or older (≥40 years) (12). Verbal consent was obtained via telephone. After providing verbal consent a study coordinator contacted the participant to confirm their willingness to participate and verify the shipping address to which a study self-testing kit was shipped and received by the participant within 24-48 hours. This kit contained a thermometer (CVS Health, Woonsocket, RI), a pulse oximeter (Zewa, Fort Myers, FL) and supplies for self-testing.(13) A study coordinator scheduled a video or phone study visit (Day 0) to occur upon receipt of the study kit to instruct participants on self-testing procedures and appropriate use of the pulse oximeter. Study visits occurred via phone for days 0, 3, 7, 14, and 21. Participants attended an in-person follow-up visit between study days 28 and 60 if they were asymptomatic at the time, consistent with local hospital infection control procedures.

Vital signs (heart rate, oxygen saturation, and temperature) were prospectively collected and recorded by participants for 14 days and reviewed with the study coordinator at each study visit. Participants were requested to call the study team and their primary care physician for oxygen saturation values <93%. At each time point participants completed a 32-item influenza Patient –Reported Outcome (FLU-PRO) (14) questionnaire verbally over the phone with the study coordinator. This questionnaire assessed participants’ sense of physical well-being including symptoms. Symptoms were reported using a Likert scale. This instrument has been previously validated for patient reported outcomes for influenza and other respiratory viruses(14-17). The FLU-PRO was modified to allow participants to report symptoms they perceived to be related to COVID-19 that were not already listed on the questionnaire (e.g. ageusia and anosmia). These additional items were not included in the mean FLU-PRO score. Study data were collected and managed using a REDCap electronic data capture tool hosted at Johns Hopkins University (18).

Preceding symptoms were recorded at enrollment and present symptoms recorded on study days. Summary statistics including prevalence, incidence, and duration were calculated for baseline demographics, baseline comorbidities and for time-varying parameters such as vital signs and modified FLU-PRO symptoms. FLU-PRO total score means and symptom domain (e.g. respiratory) means were calculated (16). To identify common symptom patterns at the onset of illness, we evaluated the frequency of combinations of CDC COVID-19 case definition symptoms in addition to diarrhea and weakness using an upset plot (19). Symptom prevalence was categorized by week of illness. To reduce the effect of recall bias, prior symptoms were included in the determination of prevalence by week of illness only if symptoms had started within a week of enrollment. To evaluate the correlation between oxygen saturation at rest and with ambulation, a Pearson’s correlation was performed, and a Bland-Altman plot was created. Kaplan-Meier plots were created to describe the time from symptom onset to a return to usual health and the time to a return to usual activities. After checking the proportional hazards assumption, Cox regression was performed to evaluate for baseline demographics and duration of illness affecting activities. Univariate negative log binomial regression was performed to evaluate the association between age, sex, baseline comorbid conditions (dichotomous), heart rate (continuous), temperature (continuous), oxygen 4saturation (continuous) on the occurrence of a subsequent hospitalization. Logistic regression was performed for significant values and an Area Under the Curve (AUROC) determined. A random effects logistic regression model was used to evaluate for an association oxygen level as a time varying covariate and subsequent hospitalization. Days since symptom onset (continuous), age (continuous) and resting oxygen saturation (continuous), and an interaction term between duration of illness and resting oxygen saturation (continuous) were candidate model covariates. Models were compared using Akaike information criterion (AIC) model estimates. Persons that withdrew or were lost to follow-up were excluded from regression analyses. The sample size includes that which was operationally obtainable during the described study period in a Master Protocol study (clinicaltrials.gov NCT04496466). Analyses were performed using Stata version 16.0 (StataCorp LLC, College Station, TX, USA) and figures were created using Stata or R statistical platform version 4.0.1 (R Foundation).

## RESULTS

From April 21 to June 23, 2020, 118 participants enrolled a median of 5.0 (3.0-10.0) days from symptoms onset, while a total of 115 others were contacted but refused participation (Figure 1). Participants were a median of 56.0 (IQR, 50.0 to 63.0) years of age, 42.4% male (N = 50), and the median Charlson Comorbidity Index was 2 (IQR, 1 to 3) (Table 1). In the prior two weeks before developing symptoms, 40.2% (N=39) had contact with someone with confirmed COVID-19 and an additional 21.9% (N=21) had contact with someone with symptoms concerning for unconfirmed COVID-19. The duration of symptoms at enrollment was 5.0 days (IQR, 3.0 to 10.0) (Table 1).

**Figure 1.**
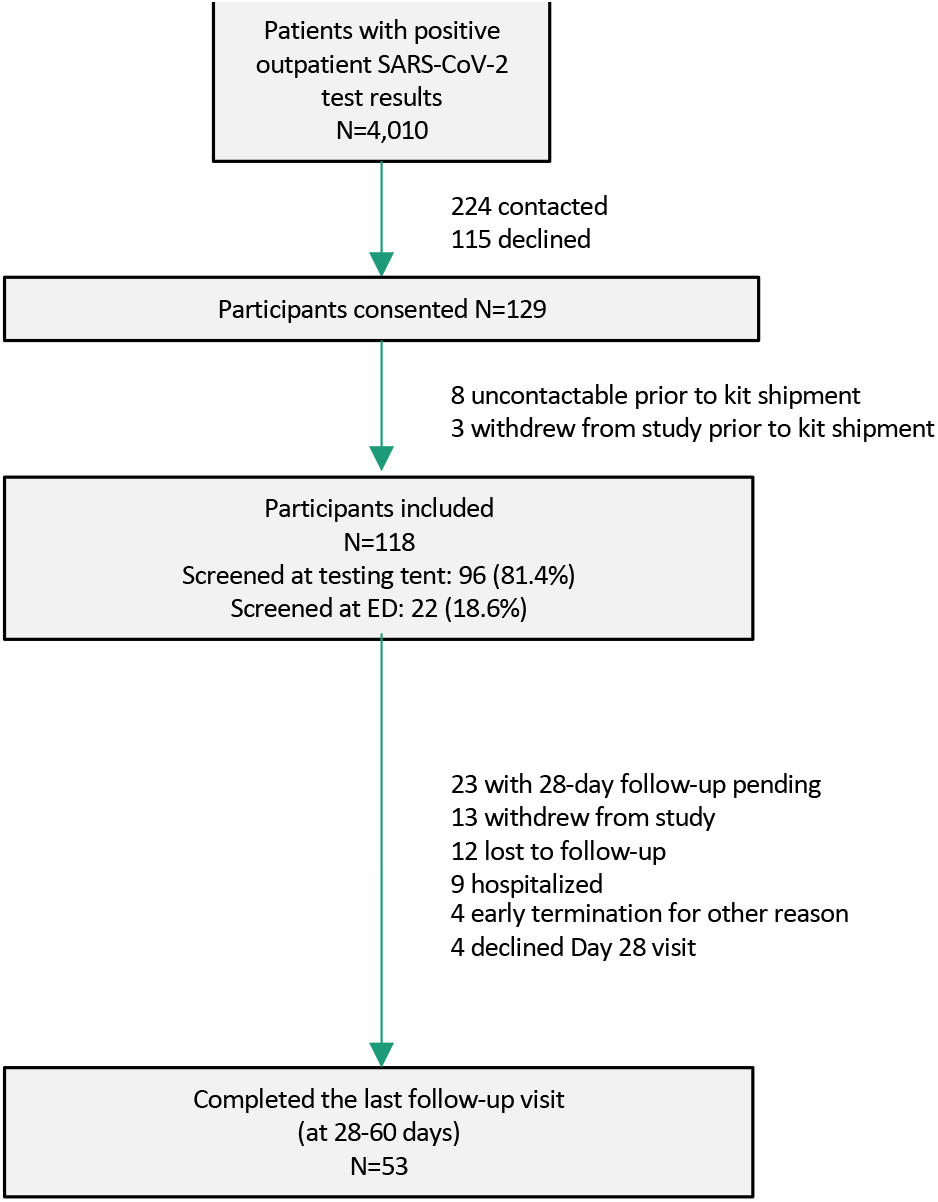
Screening, enrollments, and follow-up.

**Table 1.**
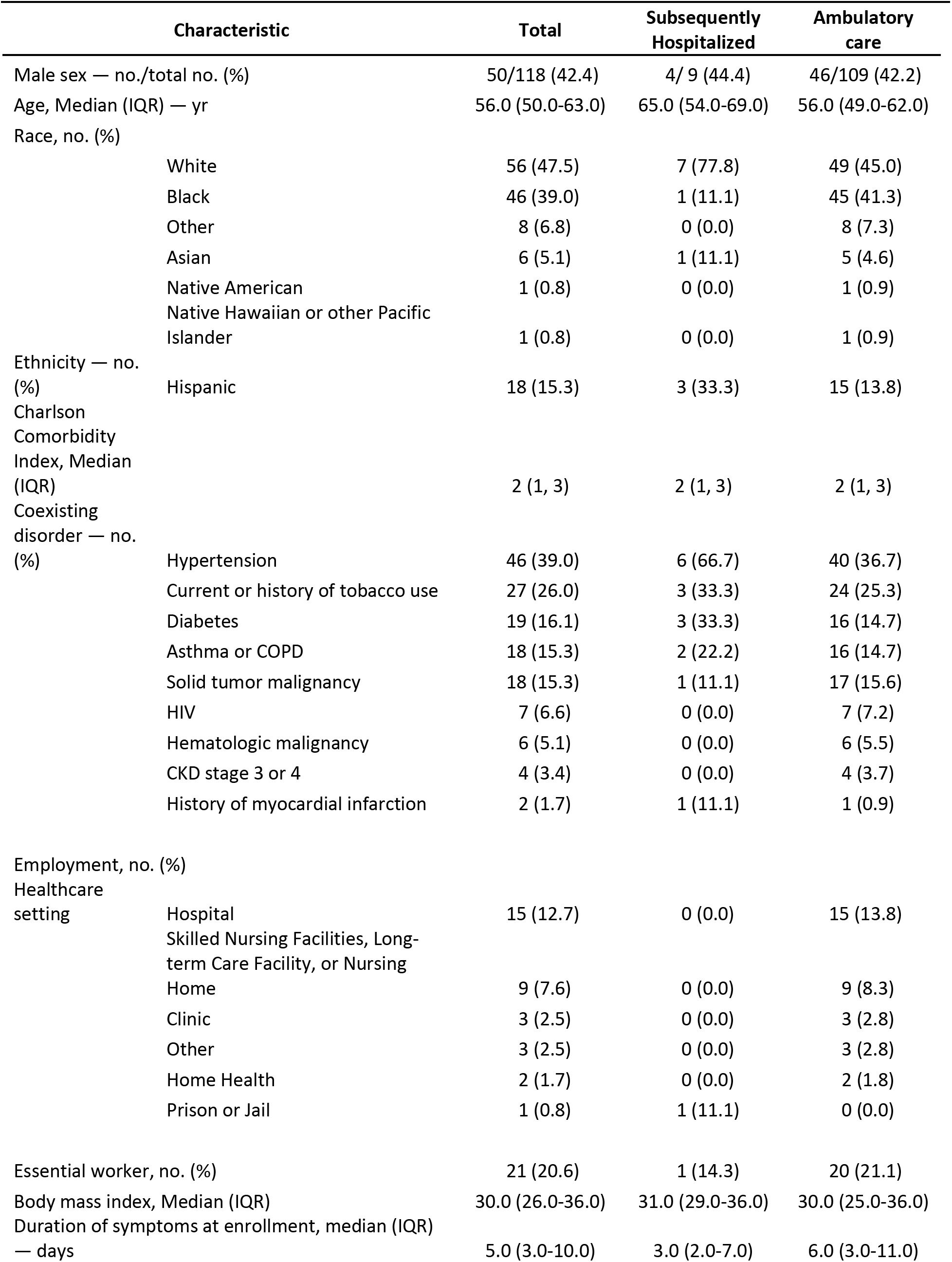
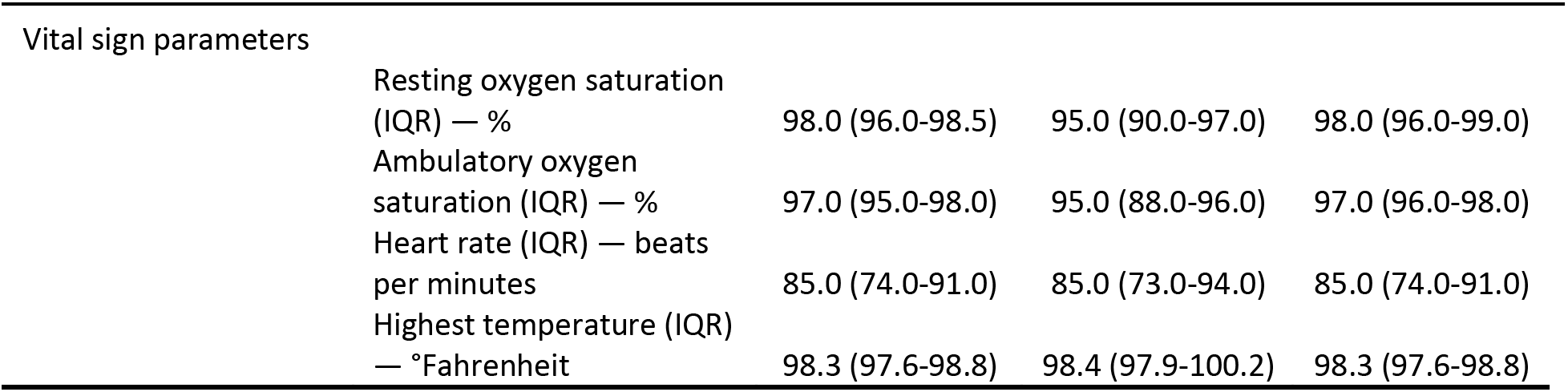
Baseline demographics and clinical characteristics of cohort stratified by subsequent hospitalization.

| Characteristic |  | Total | Subsequently Hospitalized | Ambulatory care |
| --- | --- | --- | --- | --- |
| Male sex — no./total no. (%) |  | 50/118 (42.4) | 4/ 9 (44.4) | 46/109 (42.2) |
| Age, Median (IQR) — yr |  | 56.0 (50.0-63.0) | 65.0 (54.0-69.0) | 56.0 (49.0-62.0) |
| Race, no. (%) |  |  |  |  |
|  | White | 56 (47.5) | 7 (77.8) | 49 (45.0) |
|  | Black | 46 (39.0) | 1 (11.1) | 45 (41.3) |
|  | Other | 8 (6.8) | 0 (0.0) | 8 (7.3) |
|  | Asian | 6 (5.1) | 1 (11.1) | 5 (4.6) |
|  | Native American | 1 (0.8) | 0 (0.0) | 1 (0.9) |
|  | Native Hawaiian or other Pacific Islander | 1 (0.8) | 0 (0.0) | 1 (0.9) |
| Ethnicity — no. (%) |  |  |  |  |
|  | Hispanic | 18 (15.3) | 3 (33.3) | 15 (13.8) |
| Charlson Comorbidity Index, Median (IQR) |  | 2 (1, 3) | 2 (1, 3) | 2 (1, 3) |
| Coexisting disorder — no. (%) |  |  |  |  |
|  | Hypertension | 46 (39.0) | 6 (66.7) | 40 (36.7) |
|  | Current or history of tobacco use | 27 (26.0) | 3 (33.3) | 24 (25.3) |
|  | Diabetes | 19 (16.1) | 3 (33.3) | 16 (14.7) |
|  | Asthma or COPD | 18 (15.3) | 2 (22.2) | 16 (14.7) |
|  | Solid tumor malignancy | 18 (15.3) | 1 (11.1) | 17 (15.6) |
|  | HIV | 7 (6.6) | 0 (0.0) | 7 (7.2) |
|  | Hematologic malignancy | 6 (5.1) | 0 (0.0) | 6 (5.5) |
|  | CKD stage 3 or 4 | 4 (3.4) | 0 (0.0) | 4 (3.7) |
|  | History of myocardial infarction | 2 (1.7) | 1 (11.1) | 1 (0.9) |
| Employment, no. (%) |  |  |  |  |
| Healthcare setting |  |  |  |  |
|  | Hospital | 15 (12.7) | 0 (0.0) | 15 (13.8) |
|  | Skilled Nursing Facilities, Long-term Care Facility, or Nursing Home | 9 (7.6) | 0 (0.0) | 9 (8.3) |
|  | Clinic | 3 (2.5) | 0 (0.0) | 3 (2.8) |
|  | Other | 3 (2.5) | 0 (0.0) | 3 (2.8) |
|  | Home Health | 2 (1.7) | 0 (0.0) | 2 (1.8) |
|  | Prison or Jail | 1 (0.8) | 1 (11.1) | 0 (0.0) |
| Essential worker, no. (%) |  | 21 (20.6) | 1 (14.3) | 20 (21.1) |
| Body mass index, Median (IQR) |  | 30.0 (26.0-36.0) | 31.0 (29.0-36.0) | 30.0 (25.0-36.0) |
| Duration of symptoms at enrollment, median (IQR) — days |  | 5.0 (3.0-10.0) | 3.0 (2.0-7.0) | 6.0 (3.0-11.0) |
| Vital sign parameters |  |  |  |  |
|  | Resting oxygen saturation (IQR) — % | 98.0 (96.0-98.5) | 95.0 (90.0-97.0) | 98.0 (96.0-99.0) |
|  | Ambulatory oxygen saturation (IQR) — % | 97.0 (95.0-98.0) | 95.0 (88.0-96.0) | 97.0 (96.0-98.0) |
|  | Heart rate (IQR) — beats per minutes | 85.0 (74.0-91.0) | 85.0 (73.0-94.0) | 85.0 (74.0-91.0) |
|  | Highest temperature (IQR) — °Fahrenheit | 98.3 (97.6-98.8) | 98.4 (97.9-100.2) | 98.3 (97.6-98.8) |

### Symptoms and physiologic parameters at the onset of illness

The most common initial symptoms were measured or suspected fever (28.0%), dry cough (23.7%), body aches (21.2 %), weakness or fatigue (20.3 %), and headache (17.0 %)(Appendix Figure S1). There were five (4.2%) asymptomatic participants and four patients (3.4%) who were pauci-symptomatic (one symptom prior to or at enrollment). Asymptomatic patients were tested because they had a positive contact (N=3) or during screening for medical encounters (N=2).

### Symptoms and physiologic parameters during the clinical course of disease

Among those reporting symptoms, the most common during the first week since onset included weakness/fatigue (67.3%), cough (58.0%), headache (43.8%), and sore throat (34.8%) (Appendix Table 1; Figure 2A). Repeated unexpected symptoms included a skin burning sensation (N=3) and a smell of burning wood (N=2). During the first month of illness, the prevalence of symptoms decreased but a substantial proportion of individuals still reported weakness (15.4%) or a dry cough (18.3%) (Figure 2A and 2B).

**Figure 2.**
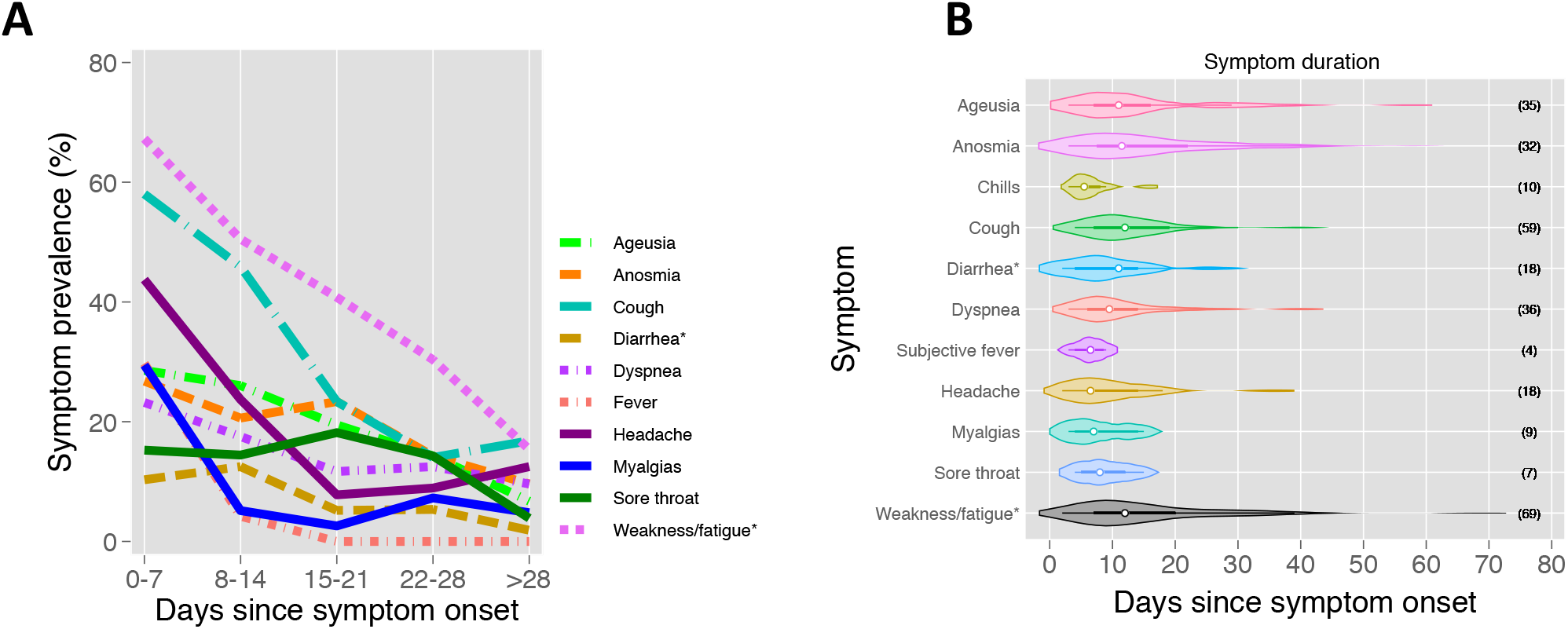
(A) Symptom prevalence by week of illness per a FLU-PRO questionnaire and additional COVID-19 specific questions and (B) violin plots of symptom duration centered by onset of the first symptom(s) to illustrate the course of disease over time from questionnaire responses during the study period *Not present in interim April 2020 CDC COVID-19 case definition.

The effect of symptoms on activities of daily living and illness severity varied greatly at any given point and over time (Figure 3). Interestingly, during the first week of illness, 43.1% of patients reported no effect of their symptoms on daily activities. Participants returned to their usual health a median of 20 days (IQR, 13 to 38) from the onset of symptoms, and the median time to returning to usual activities was 17 days (IQR, 11 to 28) from symptom onset (Figure 3A-B). Baseline factors of age, sex, or comorbid conditions were not associated with a delay in return to health or to usual activities with unadjusted Cox proportional hazards regression (data not shown). Notably, while the majority 63.7% of participant had no symptoms or only had mild symptoms during the first week of illness a substantial proportion continued to have mild or moderate symptoms for over one month (Figure 3C-E). During the third and fourth week of illness, only 52.6% and 65.5% of respondents had returned to their usual health, respectively.

**Figure 3.**
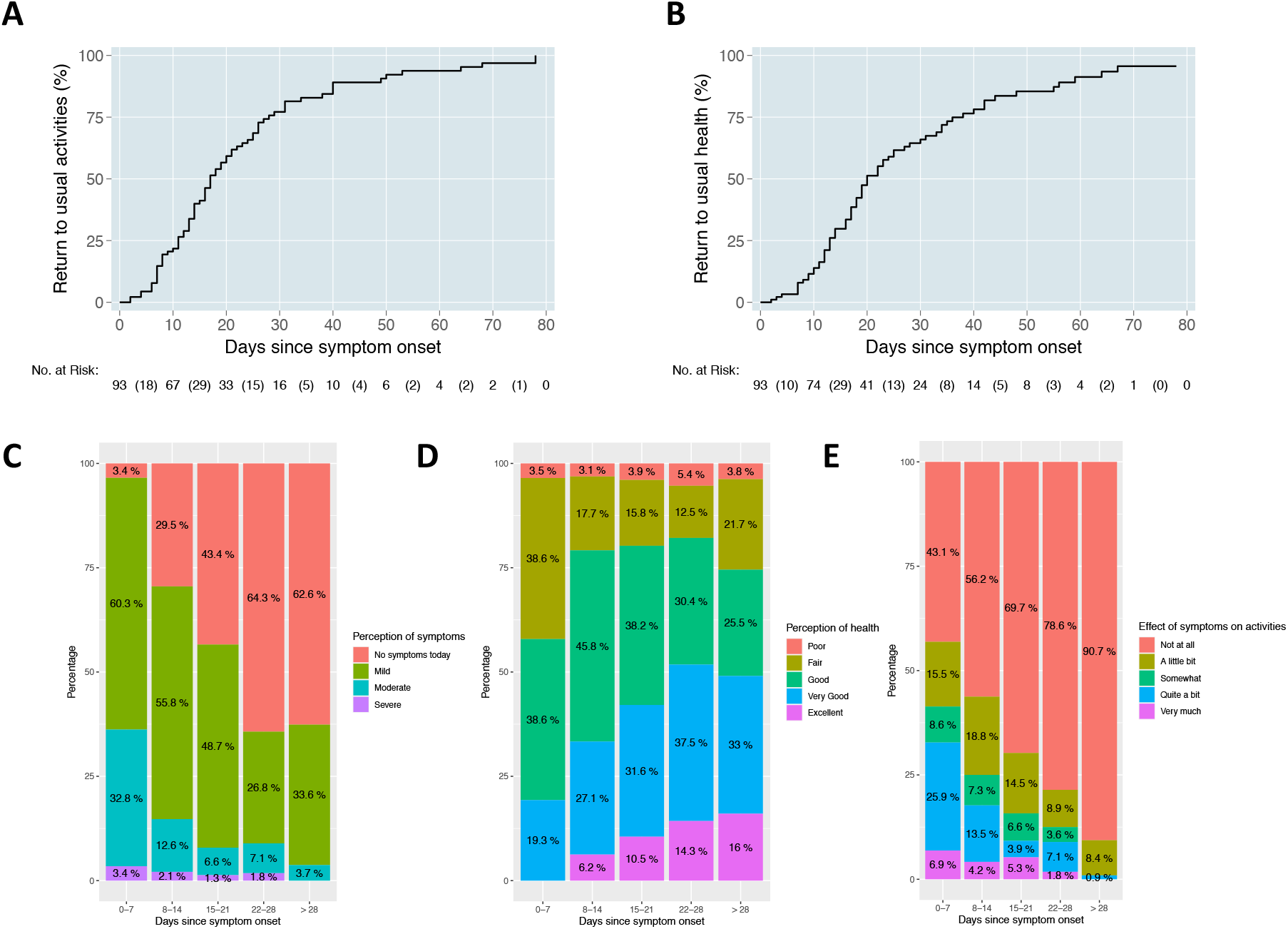
Kaplan-Meier curve of time to participants returning to usual activities (A) and to usual health (B). Severity of disease during first month of illness among those with symptomatic outpatient COVID-19 including (C) perception of disease, (D) perception of health, and (E) the effect on usual activities.

### Oxygen saturation, heart rate, temperature, and subsequent hospitalization

The initial median oxygen saturation values (SaO_2_) at rest were 98.0% (IQR, 96.0 to 98.5) and during ambulation were 97.0% (IQR, 95.0 to 98.0) (Table 1). Correlation between walking and ambulatory SaO_2_ was 0.61, with a larger difference between values noted at lower oxygen saturations (Appendix Figure S2A). Resting SaO_2_ at 92% or less occurred among 11.1% (N=8) of participants at a median 11.5 days from symptom onset (IQR, 10, 14), but only accounted for 3.4% of all SaO_2_ levels. When evaluating the mean value of respiratory symptom questions at the time of oxygen measurements, five out of eight individuals with an ambulatory SaO_2_ ≤92% and two out of five individuals with a resting SaO_2_ ≤92 had mild or no respiratory symptoms (Appendix Figure S2B-C). Additionally, there were 8 participants with at least one low (≤92%) ambulatory SaO_2_ and 4 participants with at least one low resting home SaO_2_ who did not seek medical attention, despite prior guidance.

Low oxygen saturation (<93%) was the leading factor for 5 participants being sent to the ED, followed by dyspnea (N=4), diarrhea (N=2), fever (N=1), chest pain (N=1), and elevated blood pressure (N=1). During the study period, 13 (11.0%) presented to the emergency department and 9 (7.6%) required hospitalization (Appendix Table S2). The median time from symptom onset to hospitalization was 11 days (IQR, 9 to 12).

Baseline demographics and initial study vital signs were evaluated for associations with subsequent hospitalization. Each year of age was associated with 9% increased odds of subsequent hospitalization (OR: 1.1, CI, 1.00 to 1.19, p=0.04; IRR: 1.1, CI, 1.00 to 1.17, p=0.049). Other baseline demographics or comorbidities were not associated with an increased risk of subsequent hospitalization (Appendix Figure S3).

Among those who were subsequently hospitalized, the initial median SaO_2_ at rest was 95.0 percent (IQR, 90.0 to 97.0) and during ambulation was 95.0 percent (IQR, 88.0 to 96.0) (Table 1; Figure 4A-B). Temperature, heart rate, and SaO_2_ were plotted over time and stratified by a need for subsequent hospitalization (Figure 4C-F). However, each percent decrease in value of the first recorded resting SaO_2_ (N=91) was associated with a 30.0% increased relative risk of a subsequent hospitalization (IRR: 1.3, CI: 1.413 to 1.49, p= 0.004; OR 1.7, CI: 1.18 to 2.37, p = 0.004). While the initial ambulatory SaO_2_ was similarly associated with subsequent hospitalization (OR: 1.76; CI, 1.24 to 2.48, p=0.001), temperature (OR: 1.5 per degree Fahrenheit; CI, 0.81 to 2.69, p=0.20) and heart rate (OR: 1.0 per 10 beats per minute; CI, 0.58 to 1.88, p=0.90) were not. The AUROC for the initial SaO_2_ for predicting subsequent hospitalization was 0.85 (CI, 0.71 to 0.89).

**Figure 4.**
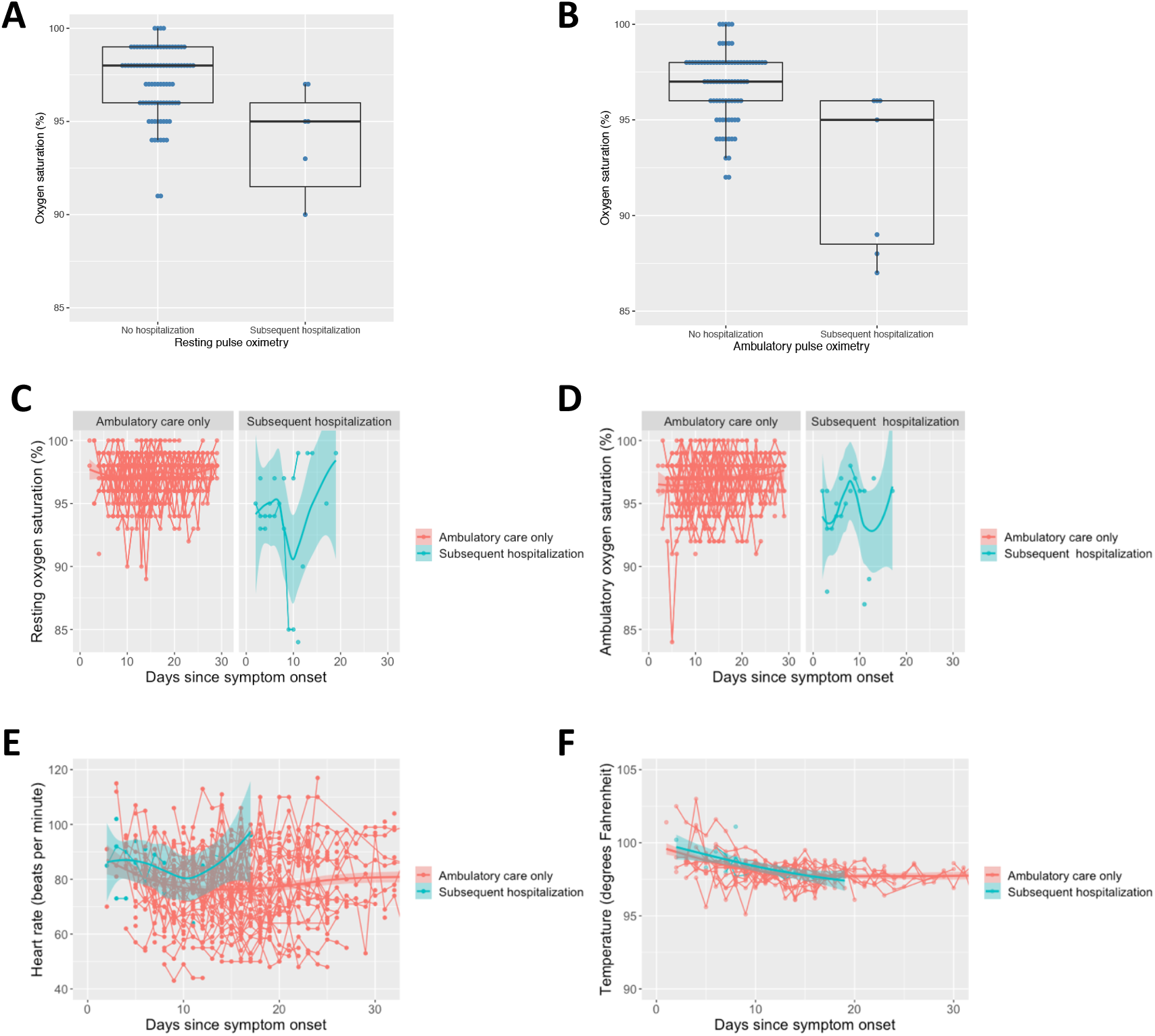
Box plots of resting (A) and ambulatory (B) oxygen saturation (%) at enrollment among those that subsequently were hospitalized and those that were not during the study period. Outpatient vital signs over time by duration of symptoms stratified by subsequent hospitalization requirement including (C) resting oxygen saturation, (D) ambulatory oxygen saturation, (E) heart rate (beats per minute), and (F) temperature (degrees Fahrenheit).

To further explore the utility of a resting SaO_2_, the performance characteristics of the individual-level nadir of resting SaO_2_ were determined for those within the first week of illness (N=42). The AUROC was found to be 0.84 (CI, 0.67 to 1.0) for predicting subsequent hospitalization. SaO_2_ values of 93% or below in the first week of illness were specific (92.1%) but not sensitive (50%) for predicting subsequent hospitalization with a 6.3 positive likelihood ratio. There was a negative likelihood ratio of zero for subsequent hospitalization among those with a resting oxygen saturation that went no lower than 96% (Appendix Table S3).

To evaluate for the association of oxygen saturation levels over time, a random effects logistic regression model was fit adjusting for age. The model was more parsimonious according to a lower AIC when including age as a covariate. Individuals at the same duration of illness were found to have a 6.6-fold increased crude odds (CI: 1.41 to 31.23, p = 0.02) and a 6.1-fold increased adjusted odds of subsequent hospitalization per every SaO_2_ percent decrease (CI: 1.41 to 31.23, p=0.02).

## DISCUSSION

This may be the first study to prospectively characterize the incidence, intensity, and duration of COVID-19 in an outpatient setting. Presenting symptoms were diverse, persistent, and uncommonly required hospitalization, especially when high ambulatory blood oxygen concentration were sustained.

These findings support early observations from hospitalized patients that symptoms may persist long-term after acute COVID-19 illness (6, 7, 20, 21). Despite outpatient COVID-19 being considered generally mild, we also found that respiratory and systemic symptoms persistent for weeks, notably longer than with common respiratory viruses(16, 17). Our findings were consistent with a cross-sectional survey that found that 35% of outpatient COVID-19 positive respondents had not returned to their usual state of health between 2 to 3 weeks from diagnosis(6). Additionally, cough, fatigue, or shortness of breath were present among 43%, 35%, and 29%, respectively, among those that initially reported symptoms(6). In comparison, participants in our cohort reported cough among 23.4%, weakness or fatigue among 40.8%, and shortness of breath among 11.7% during the third week of illness regardless of initial symptoms. Weakness or fatigue was the most pervasive symptom and almost one-third of participants reported fatigue after 22-28 days of illness. Between 10-20 percent of participants continued to have some degree of cough, headache, or anosmia as long as a month or more after the onset of symptoms. The prolonged duration of loss of taste and smell has been previously noted in the study using telephone surveys and is consistent with our prospective findings(6).

While hospitalization was uncommon in our outpatient cohort, low ambulatory or resting oxygen saturation (<93%) were predictive for requiring subsequent hospitalization, suggesting a role for home pulse oximetry in outpatient management of COVID-19. We found that the initial SaO_2_ value, a SaO_2_ nadir during the first week of illness, or any SaO_2_, adjusting for duration of symptoms, is predictive of subsequent hospitalization. We found no major difference in the diagnostic yield between resting and ambulatory SaO_2_ in this setting. The predictive value of outpatient oxygen saturation values have been previously described (5, 22). One study provided pulse oximeters to participants with COVID-19 who presented to an ED or outpatient testing centers; 29% required subsequent hospitalization. Low oxygen saturation detected on pulse oximetry was associated with hospitalization and more severe outcomes(22).

Although in the present study pulse oximetry was predictive of hospitalization, the measure alone often was not sufficient. Some persons needed hospitalization for non-respiratory symptoms (e.g. diarrhea) that logically were not detected by lower oxygen saturation. In addition, no single oxygen saturation reading alone predicted outcome as there was overlap in both the resting and ambulatory oxygen saturations of those who remained at home and those who were hospitalized. Therefore, pulse oximetry may be most useful as an adjunct to clinical monitoring of high risk populations such as those over 60 years of age, males, and persons with elevated body mass index (5, 23).

While this is the largest prospective outpatient cohort to characterize the clinical course of COVID-19, the study has several limitations. First, this study predominantly included older individuals to increase the statistical power for severe outcomes given the known association between age and hospitalization(12). Symptoms including severity and duration of illness may differ considerably in younger individuals and our results are more generalizable for persons of similar ages (24). Outpatients have been previously found to be younger and have less comorbidities compared to hospitalized patients(25). Second, the recruitment strategy may have skewed the study population. For instance, participants with milder symptoms and those with altruistic professions (e.g. healthcare workers) could have been more likely to participate in the study processes than others. A quarter of participants had symptoms of onset 10 days prior to enrollment, and therefore a proportion of participants may have been selected who had successfully passed a time window of disease severity. Additionally, due to operational requirements, Spanish-speakers were not enrolled proportionate to cases early after study initiation and individuals without active mobile phone access were not enrolled. Third, missing data from loss to follow-up or withdrawals during the study period could have skewed the longitudinal severity of results. For example, participants with milder illness could have been more likely to withdrawal the course of the study. Despite limitations, our results help elucidate the progression of outpatient COVID-19.

The prospective cohort provides additional insight into the clinical progression of outpatient COVID-19 patients, who comprise the majority of patients with SARS-CoV-2 infection. Presenting symptoms were generally diverse and often persisted longer than expected for a respiratory virus. Hospitalization occurred among 7.6% and was associated with low home SaO_2_ values, supporting the utility of pulse oximetry as a supplemental tool for remote clinical decision making. Given the diversity of manifestations of COVID-19, immunologic studies and longer-term follow-up of these patients is warranted to determine the extent of symptoms among those with persistent symptoms. There remains great uncertainty about the long-term effects of SARS-CoV-2 infection regardless of symptom severity.

## Data Availability

The data that support the findings of this study are available on request from the corresponding author, YM. The data are not publicly available to protect privacy of research participants.

## ACKNOWLEDGEMENT

We thank Dr. John Powers, Leidos Biomedical, the National Institute for Allergy and Infectious Diseases (NIAID), and the National Institutes of Health for supplying the FLU-PRO Questionnaire.

